# Sampling Statistical Errors in Big Data Research: 3 Cases of Breast Cancer Research

**DOI:** 10.1101/2021.10.07.21264601

**Authors:** Han-Jun Cho, Eui Seok Jeong

## Abstract

Breast cancer is a major cause of female death, and various big data analysis methods have been applied to breast cancer. This study lists cases in which big data analysis was applied to breast cancer research. In addition, statistics and percentages from each specific sample were proposed. However, research on the use of big data has a blind spot that relies on sample characteristics. Therefore, before sampling big data, statistical inference should be discussed more precisely through pre-examination and sample statistical errors should be reduced by professional statistical evaluation of the analysis method. In particular, the control and experimental groups should be statistically equivalent.

## Introduction

Breast cancer(BRCA) is one of the most common cancers found in women. Also, according to the results reported in the National Cancer Center for 2021, one in four people dead from breast cancer[1]. Recently on the according to the results of the research, there has been great progress in the treatment technology of breast cancer. These methods are breast cancer research using big data. In addition, with the convenience and economy of the national health insurance system, as women’s interest in breast cancer and health increases, more patients come to the hospital at an early stage and it is possible to detect it early[2]. Big data refers to the act of making data into valuable information with a specific technology or analysis tool while having the characteristics of high physical quantity and diversity of data[3]. In addition, big data analysis in the medical industry is becoming important due to the increase in medical data due to the development of the use of big data in the medical service development trend. According to IBM, 16,000 hospitals worldwide are collecting patient data, with 86,400 data being generated per patient per day[4]. In such an environment, in the case of breast cancer, which has a lower recurrence rate the earlier it is detected, it is a target disease model that can build the most effective precision medicine system in big data research[5]. Also, the use of big data health care is expected to have significant effects in cancer patient health tracking, remote patient monitoring, cost reduction and reduction of misdiagnosis rates at medical institutions, and precision medicine[6]. In this study, we report the results of analysis of recurrence characteristics using machine learning(ML) and analysis of usage behavior using data provided by the The Cancer Genome Atlas(TCGA) in USA and, Health Insurance Review & Assessment Service in Korea[7].

## Methods

### Mutation gene Big data analysis using machine learning

The Cancer Genome Atlas-BRCA provided data for 652 BRCA patients with somatic non-silent mutations and clinical information. They divided into two Disease Free/Recurred groups. To identify recurrence-related mutations, four feature selection methods (Information Gain, Chi-squared test, MRMR, Correlation) and four classifiers (Naïve Bayes, K-NN, SVM, Correlation) were used[8]. We performed 5 fold-validations to find out the efficient algorithm.

### Network analysis of hospital use behavior in breast cancer patients

The network analysis for medical utilization was conducted using Cytoscape version 3.7.2. Dataset: Health Insurance Review & Assessment Service total patient sample(HIRA-NPS-2016, 2017, HIRA-APS-2016, 2017)[9].

## Results

### Case 1. A study using machine learning and mutated genes

Usually, when a gene is used as a biomarker, a small number of genes are preferred, and the characteristic of a mutant biomarker is an objective marker that can distinguish the normal or pathological condition of a target disease and predicts the treatment response. In 652 patients, when viewed with a ratio of 589 (Disease Free): 63 (Recurred/Progressed)(supplementary table 1), a sample statistical error exists, but as shown in Figure 2 A and B, three genes are strong biomarker candidates. However, when looking at Figure 2 D, only 6 patients overlapped.

### Case 2. A study on how to apply diagnosis and algorithm using mutation feature selection in machine learning

The optimal algorithm combination was Information gain-Naïve bayes, and when diagnosis using 22-42 mutation-specific genes out of 40 genes, breast cancer can be detected early with an 88.79% probability. This problem arises because of the low proportion of relapsed patients and many non-recurring patients among all breast cancer patients in the data provided by the TCGA. When statistical errors were minimized, 144 genes were used as an appropriate number when re-experimented. The best algorithm model to be used for breast cancer diagnosis was found, but there was no significant difference from the previously reported results, and the number of genes increased as the number of genes increased. As a result of re-experiment, it was found that the number of genes using about 144 genes was rather high. In addition, as more than 500 genes were used, the diagnosis rate tended to decrease.

### Case 3. A study on analyzing medical facility usage behavior using big data network technique

Looking at hospital usage behaviors in breast cancer patients can contribute to improving medical services. but, it depending on the regional characteristics of the Republic of Korea, large medical facilities are concentrated in the capital city of Seoul, so the higher the stage of breast cancer patients, the more markedly the hospital use behavior eventually moved to Seoul. Therefore, in order to solve the hospital usage behavior, it is necessary to construct a system that enables early diagnosis in a distributed form.

## Discussion

The use of big data in cancer research is increasing day by day[17]. However, setting the sample itself is very important for big data. In the case of Figures 1, 2 and Table 1, 2 mentioned above, mutant genes that can be used as biomarkers show high predictive values of recurrence and survival rates, but in machine learning using real big data, as shown in Table 3, the appropriate number of mutant genes is determined, and The ratio and the expression amount of a particular gene are very important characteristics[18]. In addition, it is difficult to apply to cancer patient treatment because cancer patient’s hospital used behavior and network analysis techniques show the regional characteristics of each country(Figure 3 and supplementary figure 1). In addition, since it depends on the population density shown in the sample, it is difficult to apply it to improving hospital use and service in countries with low population density [19].

**Table 1.** kaplan meier values overall survival rate according to the number of genes.

| Gene number | Number of Cases, Total | Number of Events | Median Months Overall (95% CI) |
| --- | --- | --- | --- |
| 40 genes mutated | 53 | 8 | 244.91 (111.99 - NA) |
| 40 genes wild-type | 599 | 21 | NA |
| 7 genes mutated | 14 | 7 | 68.89 (44.84 - NA) |
| 7 genes wild-type | 638 | 22 | NA |
| 3 genes mutated | 6 | 4 | 93.76 (68.89 - NA) |
| 3 genes wild-type | 646 | 25 | NA |

Overall survival rate of 40, 7, and 3 recurrence-related common genes (Information gain, Chi-squared test, MRMR, Correlation). Rather, the fewer genes are used, the higher the number.

**Table 2.**
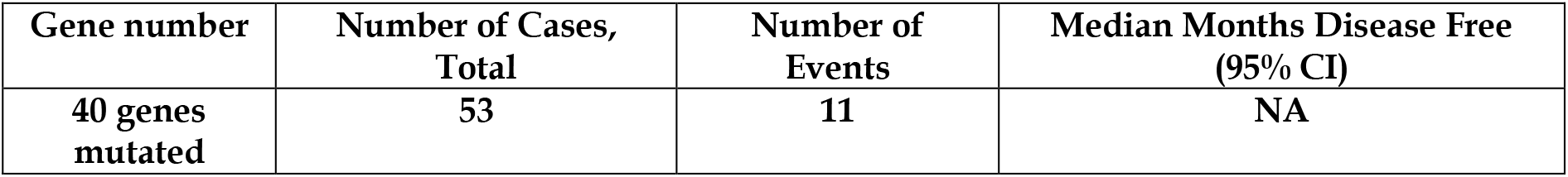
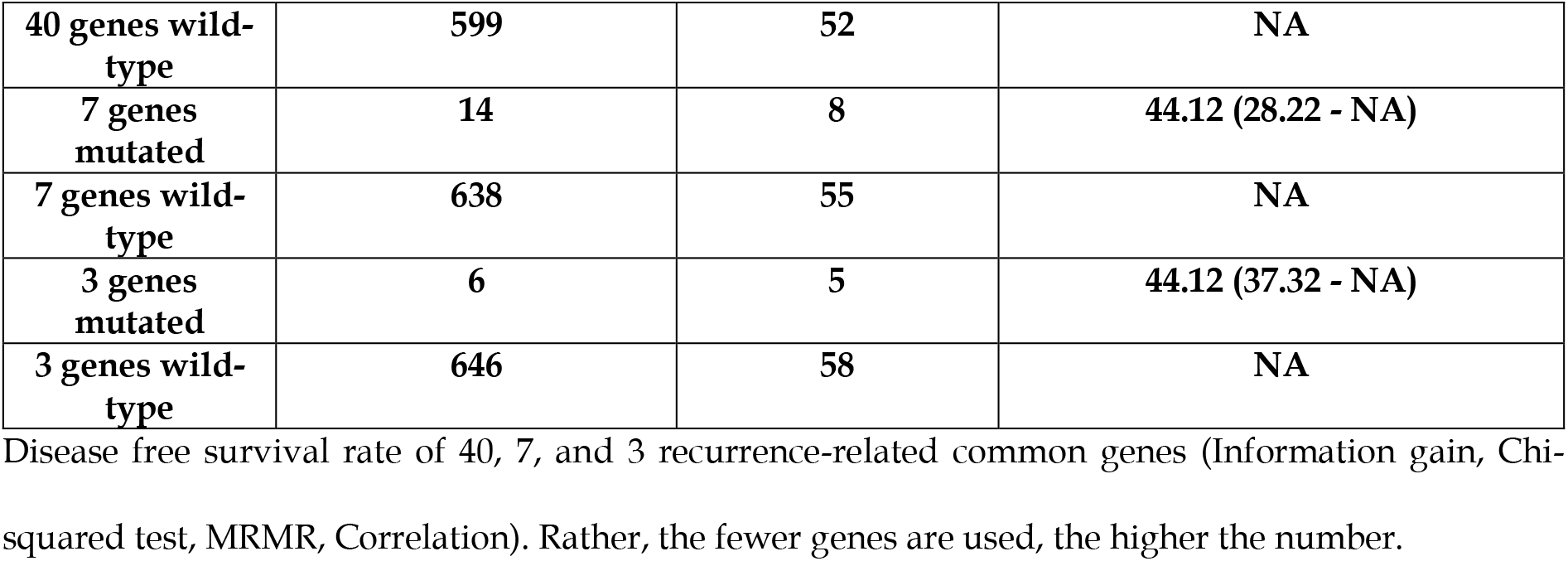
kaplan meier values disease free survival rate according to the number of genes.

| Gene number | Number of Cases, Total | Number of Events | Median Months Disease Free (95% CI) |
| --- | --- | --- | --- |
| 40 genes mutated | 53 | 11 | NA |
| 40 genes wild-type | 599 | 52 | NA |
| 7 genes mutated | 14 | 8 | 44.12 (28.22 - NA) |
| 7 genes wild-type | 638 | 55 | NA |
| 3 genes mutated | 6 | 5 | 44.12 (37.32 - NA) |
| 3 genes wild-type | 646 | 58 | NA |

Disease free survival rate of 40, 7, and 3 recurrence-related common genes (Information gain, Chi-squared test, MRMR, Correlation). Rather, the fewer genes are used, the higher the number.

**Table 3.** Optimization of 1-500(Option: random selection number) Derivation of Genetic Equivalence for Machine learning gene titration.

| BRCA Information gain-Naïve bayes (Reccurence 0-1) |  |  |  |  |  |
| --- | --- | --- | --- | --- | --- |
| K | Accuracy | Precision | Recall | Classification error | Correlation |
| 1 | 87.61% | 43.80% | 50.00% | 12.39% | 0.00% |
| 2 | 87.95% | 93.95% | 51.37% | 12.05% | 15.52% |
| 3 | 88.12% | 94.03% | 52.05% | 11.88% | 19.02% |
| 6 | 88.29% | 81.73% | 53.92% | 11.71% | 22.29% |
| 12 | 88.96% | 86.21% | 56.66% | 11.04% | 31.05% |
| 22 | 88.96% | 82.18% | 57.83% | 11.04% | 31.75% |
| 42 | 86.25% | 64.81% | 58.63% | 13.75% | 22.61% |
| 77 | 88.96% | 78.29% | 60.18% | 11.04% | 33.95% |
| 144 | 90.32% | 90.17% | 62.13% | 9.68% | 44.16% |
| 269 | 87.44% | 70.33% | 66.37% | 12.56% | 36.49% |
| 500 | 87.78% | 71.50% | 68.91% | 12.22% | 40.33% |

Each value is the same as the number of appropriate biomarkers from 1 to 500, but there is an error in the possibility of a diagnosis rate because the equivalence between the control group and the experimental group of the total number of samples applied here is different.

**Figure 1.**
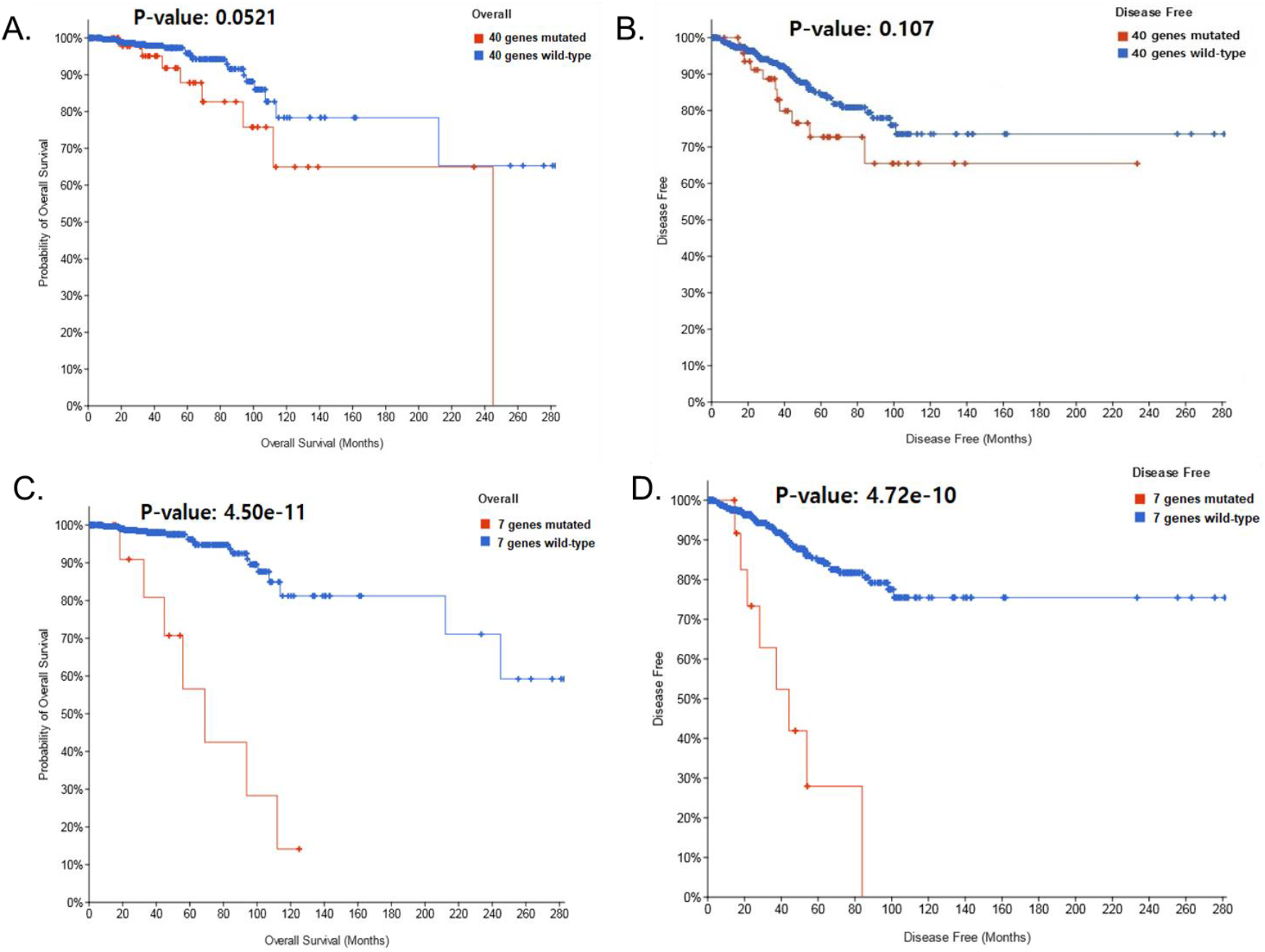
Total 7 Recurrence of Kaplan-meier specific from Recurrence-related genes. Among the 40 genes extracted by machine learning, 7 genes(ACSF3, ARID3B, KHSRP, LUZP2, RPL18A, TPI1, VWA5B2)(supplementary figure 2, 3, 4, 5, 6, 7, 8) highly related to breast cancer patients(supplementary table 2-1, 2-2). According to Kaplan Meyer statistics, the recurrence and survival prediction rates that would be expressed in all 7 specific mutant genes were rather closely related to the survival rate. In 40 vs 7, rather than using 7 genes, the predicted value is higher. The ACSF3 gene encodes a member of the acyl-CoA synthetase family that activates fatty acids by catalyzing the formation of thioester bonds between fatty acids and coenzyme A[10]. The ARID3B gene encodes a member of the ARID (AT-rich interaction domain) family of DNA binding proteins[11]. The KHSRP gene encodes a multifunctional RNA-binding protein involved in a variety of cellular processes including transcription, alternative pre-mRNA splicing, and mRNA localization[12]. The LUZP2 gene encodes a leucine zipper protein. This protein is deleted in some patients with Wilms’ tumor-Aniridia-Genitourial ornormal-mental retardation (WAGR) syndrome. Alternate splicing results in multiple script variants[13]. The RPL18A gene encodes a member of the L18AE family of ribosomal proteins, which is a component of the 60S subunit[14]. The TPI1 gene encodes an enzyme composed of two identical proteins that catalyzes the isomerization of glyceraldehyde 3-phosphate (G3P) and dihydroxy-acetone phosphate (DHAP) in glycolysis and gluconeogenesis[15]. Von Willebrand Factor A Domain Containing 5B2 (VWA5B2) is a protein-coding gene. An important paralog of this gene is VWA5B1[16]. It is inferred that the commonality of the genes is related to the process of fat synthesis. By inferring that most of the components of breasts in the human body are lipids, this is a possible decision. When 40 genes were used, the OS (overall survival) P-value: 0.0521 and DFS (Disease free survival) P-value: 0.107 as shown in Figure 1A, B P-value came out, but unlike this, all 7 genes as shown in Figure 1C, D that P-values were all significant.

**Figure 2.**
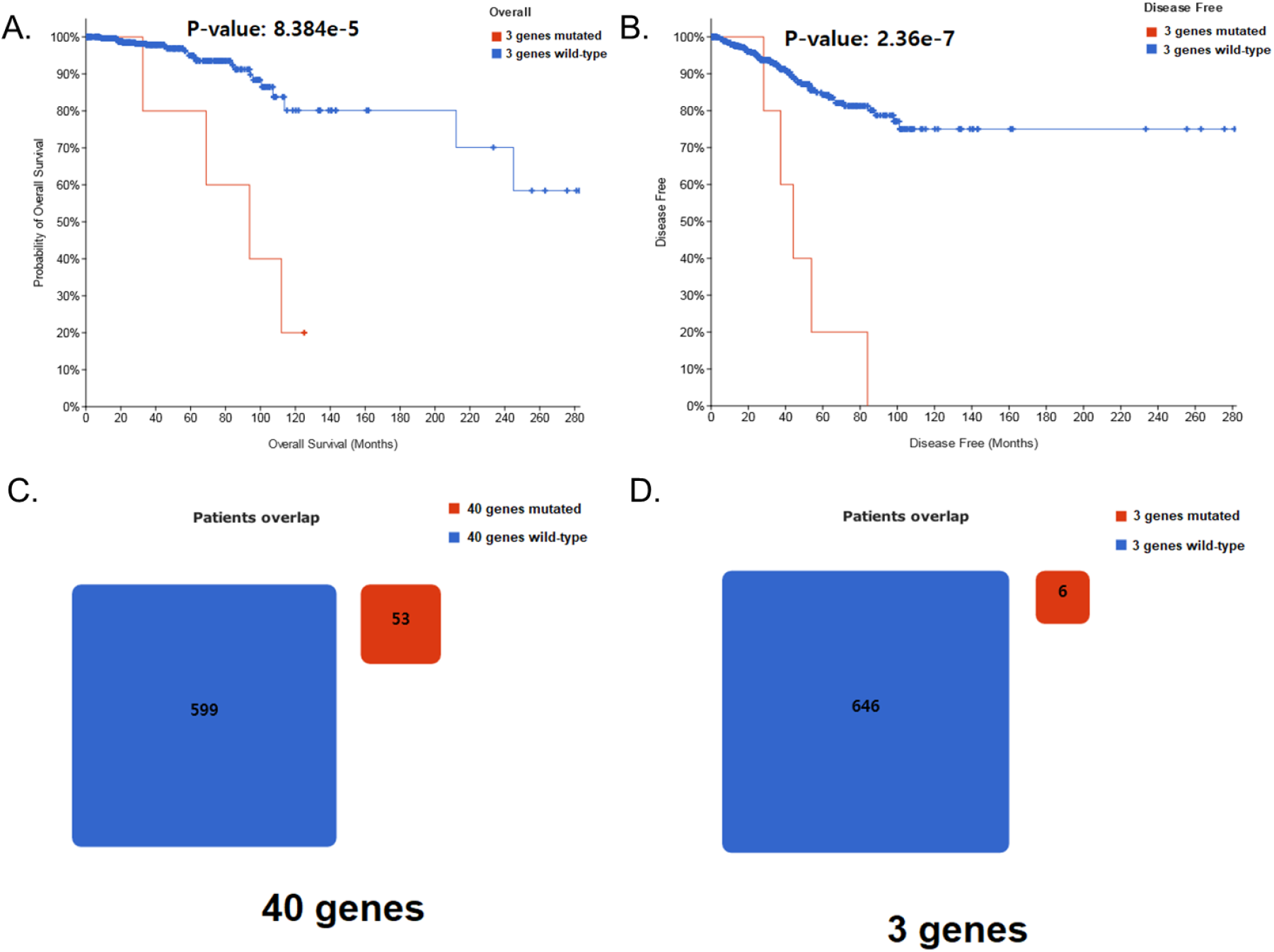
Kaplan-meier rates(OS and DFS) for 3 survival-specific genes from 4 feature selection methods. The Kaplan Meyer curve, which measured the expression rate of mutations in breast cancer patients with 3 (KHSRP, LUZP2, VWA5B2) genes, showed a very high predictive rate. This indicates that it can be easier, and only three genes can predict the stage of breast cancer patients.

**Figure 3.**
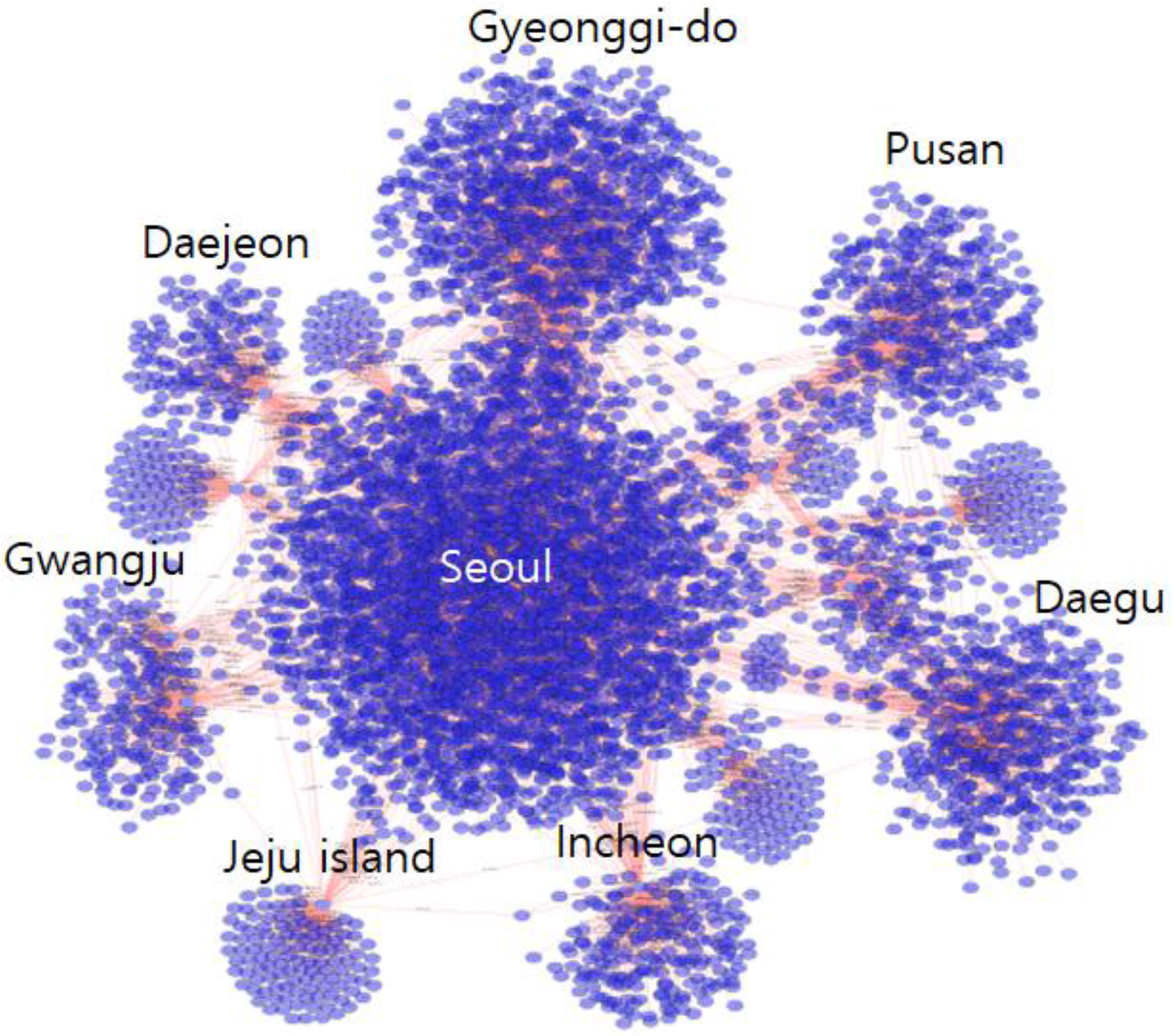
The regional distribution of BRCA patients and hospitals, its network. 19% of total hospitals are located in Seoul, whiles 42% of total breast cancer patients visit hospitals in Seoul. This indicates strong seoul-centerism. Busan, Incheon, Daegu provinces also have higher patient visits compared to the percentage of hospitals(supplementary figure 1). It all indicates metropolitan cities’ domination in hospital utilization. The network indicates that only few metropolitan cities attract most of the breast cancer patients.

It is good to try to utilize the big data that accumulates every day, but it is necessary to balance the data in order to be used in cancer research. In other words, even when using big data, the more data is used, the lower the accuracy, and the closer to disorder. However, filtering reduces the reliability of the data because the total amount in the sample is reduced[20]. Because there is such a prisoner’s dilemma as the Nash equilibrium of big data research, when using big data, it is necessary to reset the sample that is combined with the ratio of the sample rather than the simple population being formed by the organization that provides it[21-22].

In order to overcome these shortcomings of research using big data, first, the use of cancer patient data must be openly open and a clear sample range must be established. Second, deviating from the research methodology, each journal needs an evaluation team to evaluate whether the use of big data is the right analysis method. Third, it was the utilization of big data increases, it can be applied to various fields, so essential big data education of experts in various fields is required.

## Conclusion

In this study, genetic defects were important in the study of mutant genes using machine learning in breast cancer. In addition, when looking at the results of patients’ hospital use behavior through network analysis, various studies using big data are possible. However, uncertainty in the data remains. An out-of-balance especially in the proportions of the sample warns of the danger. The statistic that supports the result is very important, but it can be a statistic that applies only to a specific sample. In addition, most importantly, a clear regulation is needed to maintain the equivalence of the experimental group and the control group in the sampling of big data research.

## Data Availability

I checked the contents related to this.

https://www.cbioportal.org

https://opendata.hira.or.kr/home.do

## Acknowledge

The authors thank Dong Hyeon Lee, Da Hyun Song, You Jeong Hong and Young Geon Ji for their technical assistance with collection of data. I wrote a thesis on a completely different topic than my initial hypothesis. It is difficult to give author permission because the content of the research report is completely different. I also thank Eui-Seok Jung, who inspired us to write this paper. Even now I miss him. I pray for the repose of the deceased.

