## Supplementary for "Sampling Statistical Errors in Big Data Research: 3 Cases of Breast Cancer Research"

Table 1. Recurrence groups of Breast cancer patients from TCGA data set (N=652)

| Recurrence | Patients (%) |
|---|---|
| Disease Free(R0) | 589 (90.3%) |
| Recurred/Progressed (R1) | 63 (9.7%) |

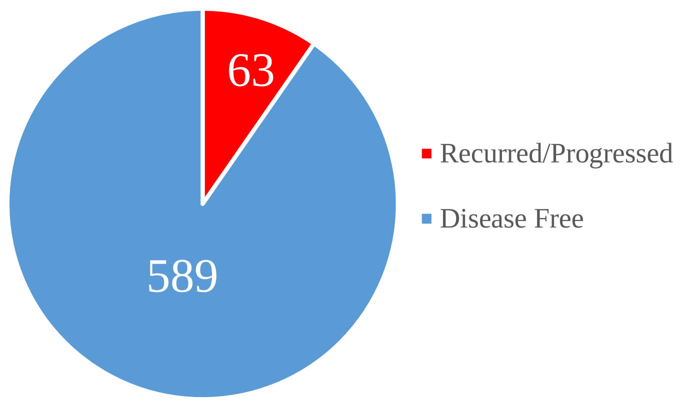


**Total of 40 recurrence-related genes were selected by ML, and of which 7 genes were identified as recurrence-specific. Each feature selection methods and performances of classifiers were useful according to disease free/recurred groups. Extremely useful genes were extracted.**

Table 2-1. Recurrence-specific 3 out of 40 common genes from top 100 genes, respectively selected by 4 feature selection methods.

| Type / Gene | Recurrence group | | Recurrence (%) | Mutation Number | Mutation/652 (%) | Mutation Type | | | Fisher's exact (p-value) | cytoband |
|---|---|---|---|---|---|---|---|---|---|---|
| | R0 | R1 | | | | Truncating | Missense (unknown) | Inframe | | |
| ACACA | 3 | 0 | 0.00% | 3 | 0.46% | 1 | 2 | 0 | 1 | 17q12 |
| ACSF3 | 2 | 1 | 33.33% | 3 | 0.46% | 1 | 2 | 0 | 0.263 | 16q24.3 |
| ARID3B | 1 | 1 | 50.00% | 2 | 0.31% | 0 | 2 | 0 | 0.184 | 15q24.1 |
| ATP5PB | 1 | 0 | 0.00% | 1 | 0.15% | 0 | 1 | 0 | 1 | 1p13.2 |
| ADGRB2 | 3 | 0 | 0.00% | 3 | 0.46% | 1 | 1 | 1 | 1 | 1p35.2 |
| C11ORF24 | 2 | 0 | 0.00% | 2 | 0.31% | 0 | 2 | 0 | 1 | 11q13.2 |
| MAJIN | 1 | 0 | 0.00% | 1 | 0.15% | 0 | 1 | 0 | 1 | 11q13.1 |
| CALCRL | 1 | 0 | 0.00% | 1 | 0.15% | 0 | 1 | 0 | 1 | 2q32.1 |
| CELSR1 | 5 | 1 | 16.67% | 6 | 0.92% | 2 | 4 | 0 | 0.458 | 22q13.31 |
| CWC25 | 2 | 0 | 0.00% | 2 | 0.31% | 0 | 2 | 0 | 1 | 17q12 |
| DNMT3B | 2 | 0 | 0.00% | 2 | 0.31% | 0 | 2 | 0 | 1 | 20q11.21 |
| ECM2 | 2 | 1 | 33.33% | 3 | 0.46% | 0 | 2 | 1 | 0.263 | 9q22.31 |
| EFCAB12 | 1 | 0 | 0.00% | 1 | 0.15% | 0 | 1 | 0 | 1 | 3q21.3 |
| ERO1A | 1 | 0 | 0.00% | 1 | 0.15% | 0 | 1 | 0 | 1 | 14q22.1 |
| EYA3 | 2 | 0 | 0.00% | 2 | 0.31% | 0 | 2 | 0 | 1 | 1p35.3 |
| FRG1 | 5 | 1 | 16.67% | 6 | 0.92% | 2 | 4 | 0 | 0.458 | 4q35.2 |
| GUCY1B1 | 2 | 0 | 0.00% | 2 | 0.31% | 1 | 1 | 0 | 1 | 4q32.1 |
| HAVCR1 | 1 | 0 | 0.00% | 1 | 0.15% | 0 | 1 | 0 | 1 | 5q33.3 |
| HK3 | 1 | 1 | 50.00% | 2 | 0.31% | 0 | 2 | 0 | 0.184 | 5q35.2 |
| HTR1E | 2 | 0 | 0.00% | 2 | 0.31% | 1 | 1 | 0 | 1 | 6q14.3 |

Table 2-2. Recurrence-specific 3 out of 40 common genes from top 100 genes, respectively selected by 4 feature selection methods.

| Type / Gene | Recurrence group | | Recurrence (%) | Mutation Number | Mutation/652 (%) | Mutation Type | | | Fisher's exact (p-value) | cytoband |
|---|---|---|---|---|---|---|---|---|---|---|
| | R0 | R1 | | | | Truncating | Missense (unknown) | Inframe | | |
| HYLS1 | 2 | 0 | 0.00% | 2 | 0.31% | 0 | 2 | 0 | 1 | 11q24.2 |
| KHDRBS2 | 1 | 0 | 0.00% | 1 | 0.15% | 0 | 1 | 0 | 1 | 6q11.1 |
| KHSRP | 0 | 2 | 100.00% | 2 | 0.31% | 1 | 1 | 0 | 0.009 | 19p13.3 |
| LILRA5 | 1 | 0 | 0.00% | 1 | 0.15% | 0 | 1 | 0 | 1 | 19q13.42 |
| LUZP2 | 0 | 2 | 100.00% | 2 | 0.31% | 0 | 2 | 0 | 0.009 | 11p14.3 |
| MAP3K4 | 2 | 0 | 0.00% | 2 | 0.31% | 2 | 0 | 0 | 1 | 6q26 |
| NDFIP2 | 2 | 1 | 33.33% | 3 | 0.46% | 0 | 3 | 0 | 0.263 | 13q31.1 |
| PDZD8 | 1 | 1 | 50.00% | 2 | 0.31% | 0 | 2 | 0 | 0.184 | 10q25.3-q26.11 |
| PITX2 | 1 | 0 | 0.00% | 1 | 0.15% | 0 | 1 | 0 | 1 | 4q25 |
| RPL18A | 2 | 1 | 33.33% | 3 | 0.46% | 1 | 2 | 0 | 0.263 | 19p13.11 |
| SIAH1 | 2 | 0 | 0.00% | 2 | 0.31% | 1 | 1 | 0 | 1 | 16q12.1 |
| SLC15A1 | 1 | 0 | 0.00% | 1 | 0.15% | 0 | 1 | 0 | 1 | 13q32.2-q32.3 |
| SMCR8 | 2 | 0 | 0.00% | 2 | 0.31% | 0 | 2 | 0 | 1 | 17p11.2 |
| SYNPR | 1 | 1 | 50.00% | 2 | 0.31% | 1 | 1 | 0 | 0.184 | 3p14.2 |
| TPI1 | 0 | 1 | 100.00% | 1 | 0.15% | 1 | 0 | 0 | 0.097 | 12p13.31 |
| TPPP2 | 2 | 0 | 0.00% | 2 | 0.31% | 0 | 2 | 0 | 1 | 14q11.2 |
| TTLL13P | 1 | 0 | 0.00% | 1 | 0.15% | 0 | 1 | 0 | 1 | 15q26.1 |
| UBE3B | 2 | 0 | 0.00% | 2 | 0.31% | 0 | 2 | 0 | 1 | 12q24.11 |
| VWA5B2 | 1 | 2 | 66.67% | 3 | 0.46% | 1 | 2 | 0 | 0.026 | 3q27.1 |
| ZNF615 | 2 | 0 | 0.00% | 2 | 0.31% | 1 | 1 | 0 | 1 | 19q13.41 |

Table 3. Recurrence data of 7 Kaplan meier specific genes from 40 Recurrence-related genes.

| Survival / Gene | Overall Survival (P-value) | Disease Free Survival (P-value) | note |
|---|---|---|---|
| ACSF3 | 0.00241 | 0.094 | **7 Genes / Total 40 Genes**<br>: Specific Gene (P-value < 0.05) |
| ARID3B | 6.27E-05 | 0.12 | |
| KHSRP | 0.269 | 0.0001616 | |
| LUZP2 | 4.82E-04 | 0.00208 | |
| RPL18A | 4.61E-07 | 0.0032 | |
| TPI1 | 0 | 2.49E-09 | |
| VWA5B2 | 3.33E-05 | 0.00179 | |

### Figure 1. Distribution of patients by medical institution type

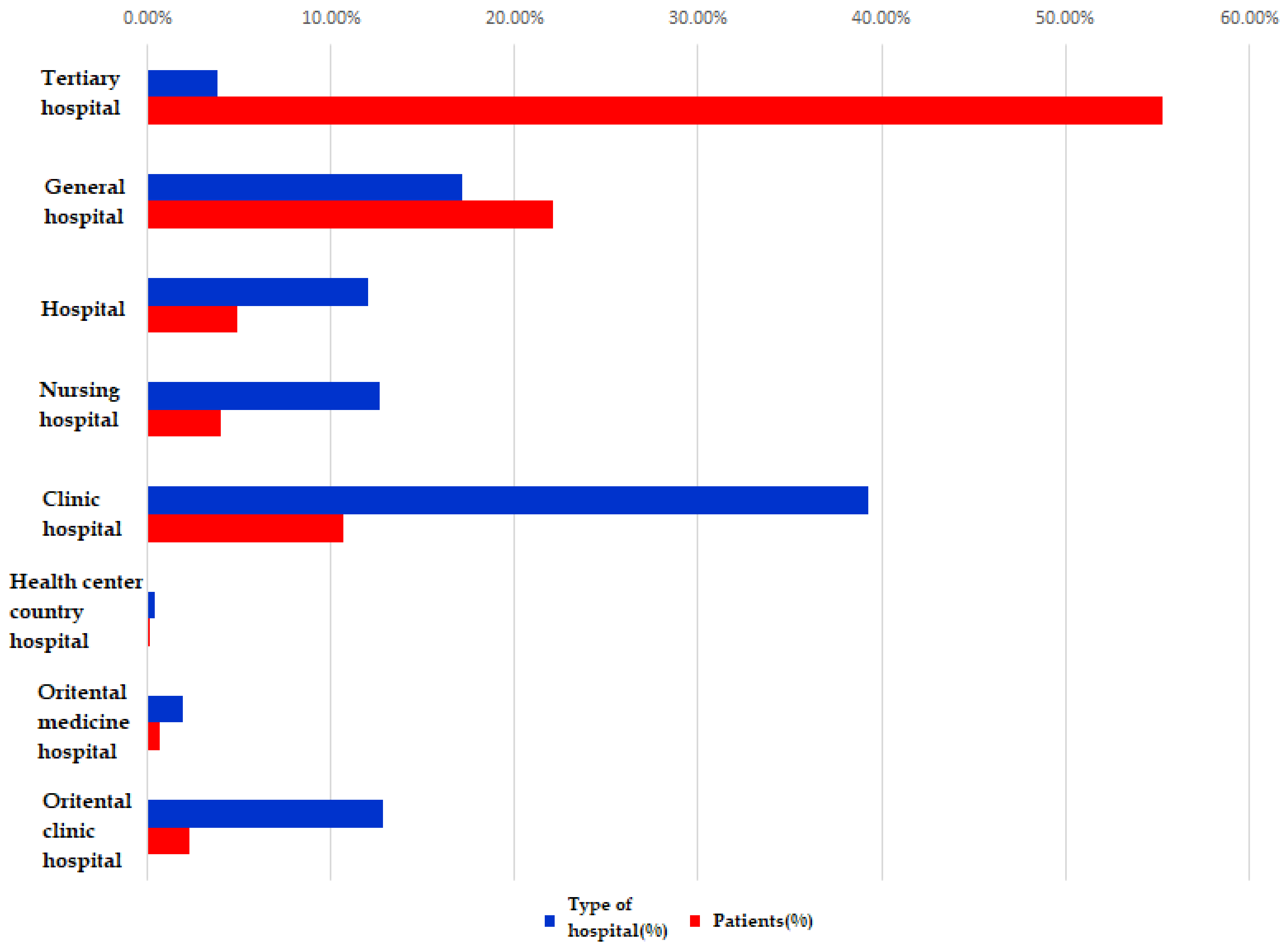

### Figure 2. KM-Curve: ACSF3 survival specific mutation gene

ACSF3

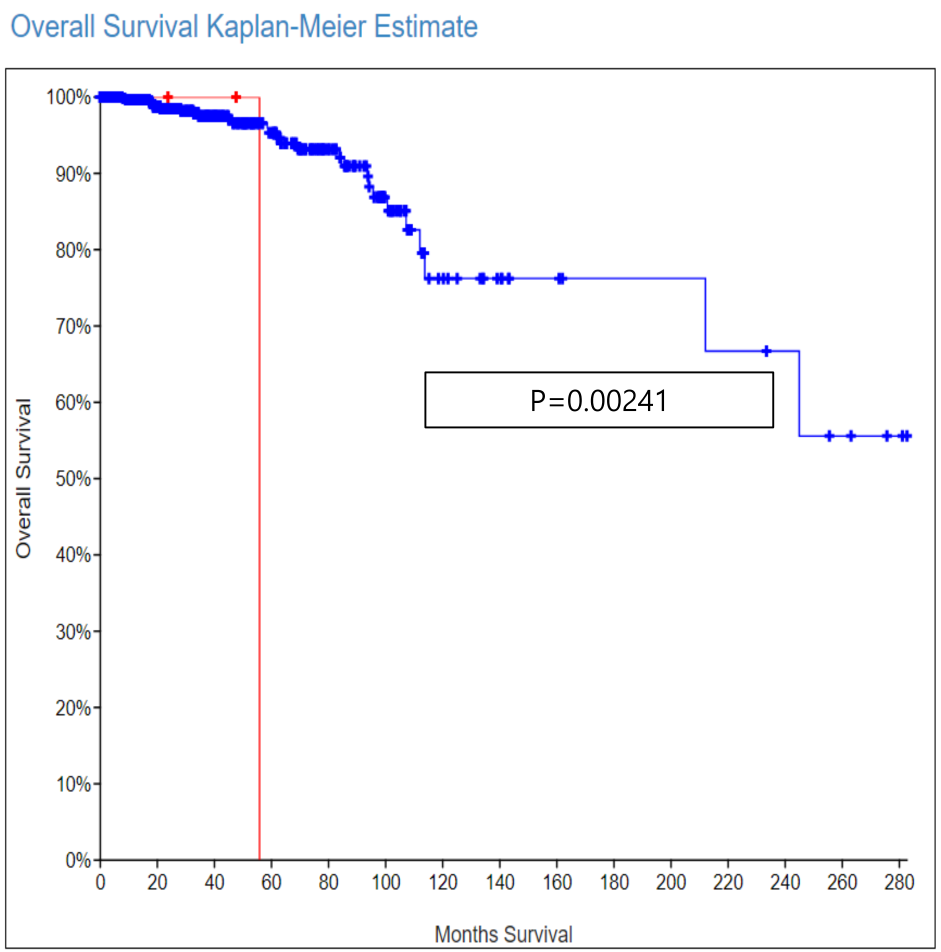


| | Number of Cases, Total | Number of Cases, Deceased | Median Months Survival |
|---|---|---|---|
| Cases with Alteration(s) in Query Gene(s) | 3 | 1 | 55.81 |
| Cases without Alteration(s) in Query Gene(s) | 649 | 28 | NA |

### Figure 3. KM-Curve: ARID3B survival specific mutation gene

ARID3B

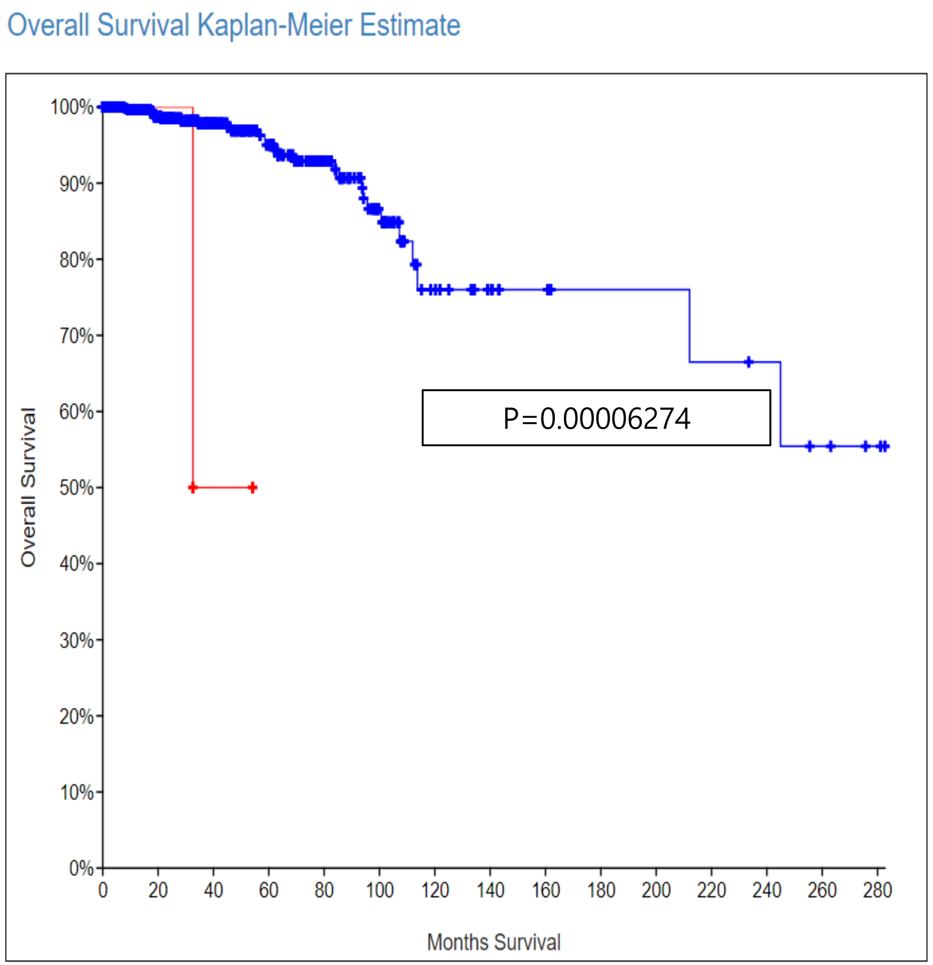


| | Number of Cases, Total | Number of Cases, Deceased | Median Months Survival |
|---|---|---|---|
| Cases with Alteration(s) in Query Gene(s) | 2 | 1 | 32.56 |
| Cases without Alteration(s) in Query Gene(s) | 650 | 28 | NA |

#### Figure 4. KM-Curve: KHSRP survival specific mutation gene

**KHSRP**

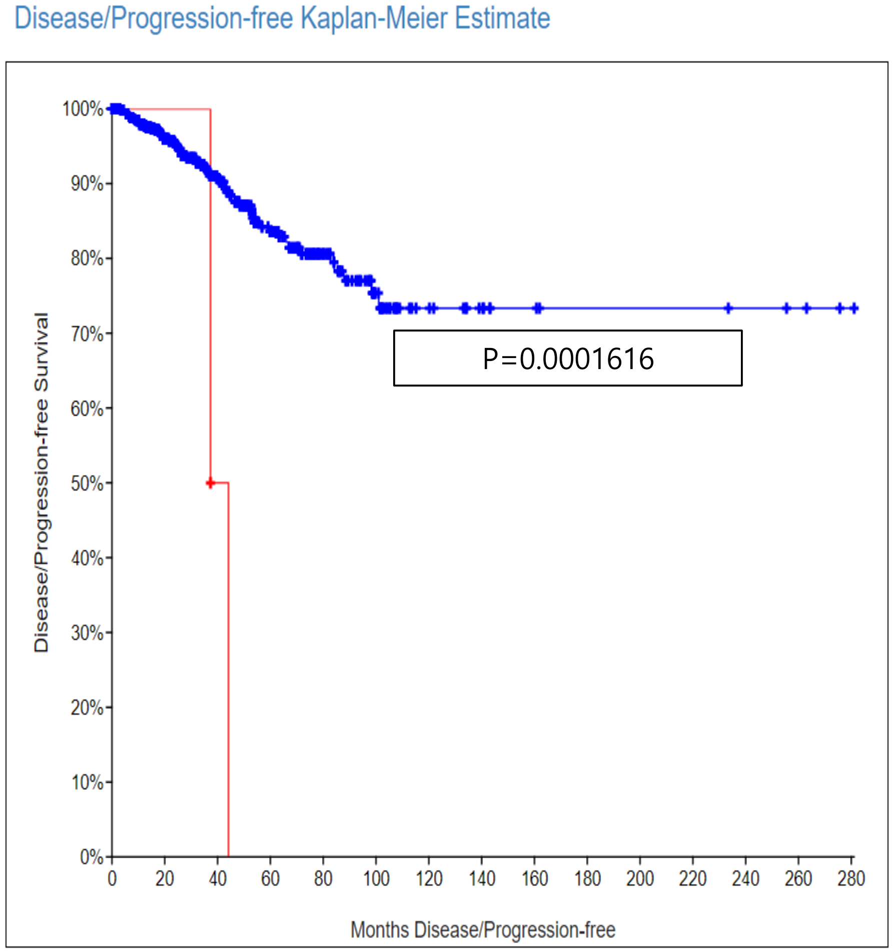


| | Number of Cases, Total | Number of Cases, Relapsed/Progressed | Median Months Disease-free |
|---|---|---|---|
| Cases with Alteration(s) in Query Gene(s) | 2 | 2 | 37.32 |
| Cases without Alteration(s) in Query Gene(s) | 650 | 61 | NA |

#### Figure 5. KM-Curve: LUZP2 survival specific mutation gene

**LUZP2**

A Overall Survival Kaplan-Meier Estimate

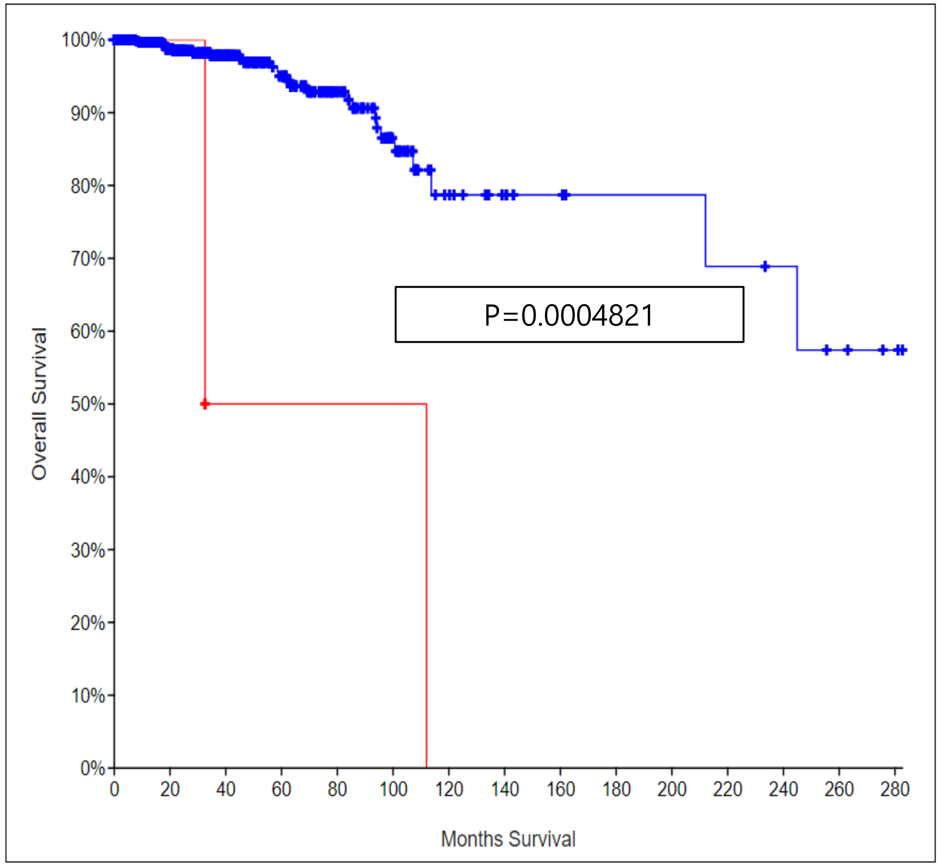


B Disease/Progression-free Kaplan-Meier Estimate

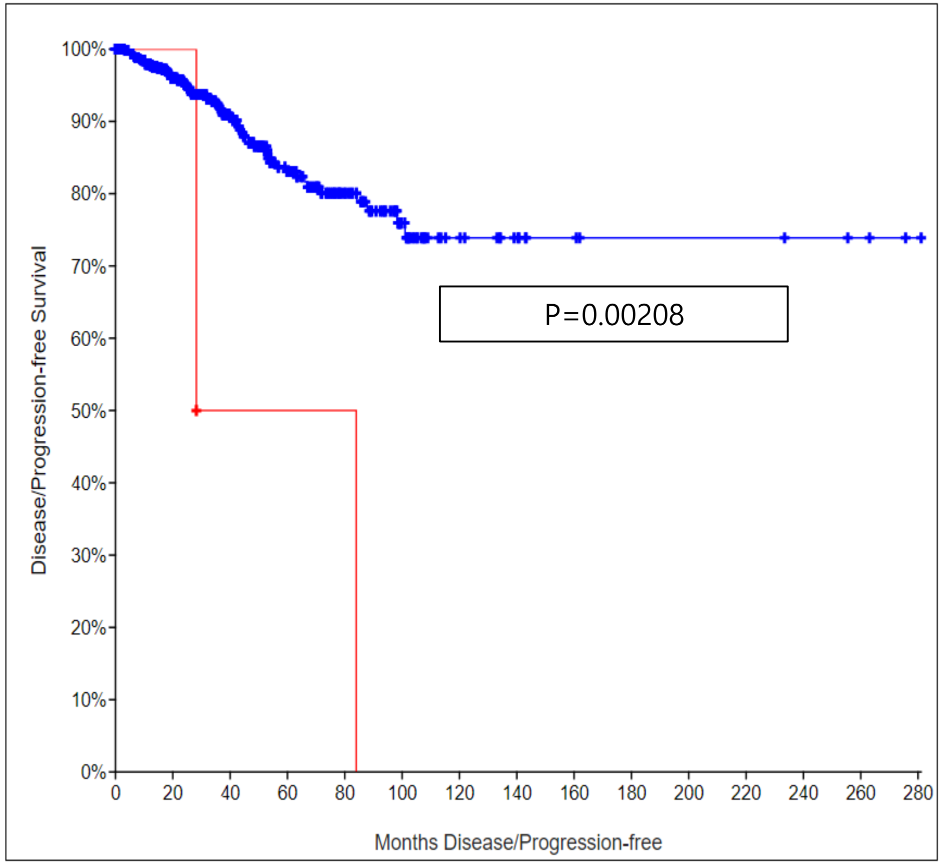


| | Number of Cases, Total | Number of Cases, Deceased | Median Months Survival |
|---|---|---|---|
| Cases with Alteration(s) in Query Gene(s) | 2 | 2 | 32.56 |
| Cases without Alteration(s) in Query Gene(s) | 650 | 27 | NA |

| | Number of Cases, Total | Number of Cases, Relapsed/Progressed | Median Months Disease-free |
|---|---|---|---|
| Cases with Alteration(s) in Query Gene(s) | 2 | 2 | 28.22 |
| Cases without Alteration(s) in Query Gene(s) | 650 | 61 | NA |

#### Figure 6. KM-Curve: RPL18A survival specific mutation gene

**RPL18A**

A Overall Survival Kaplan-Meier Estimate

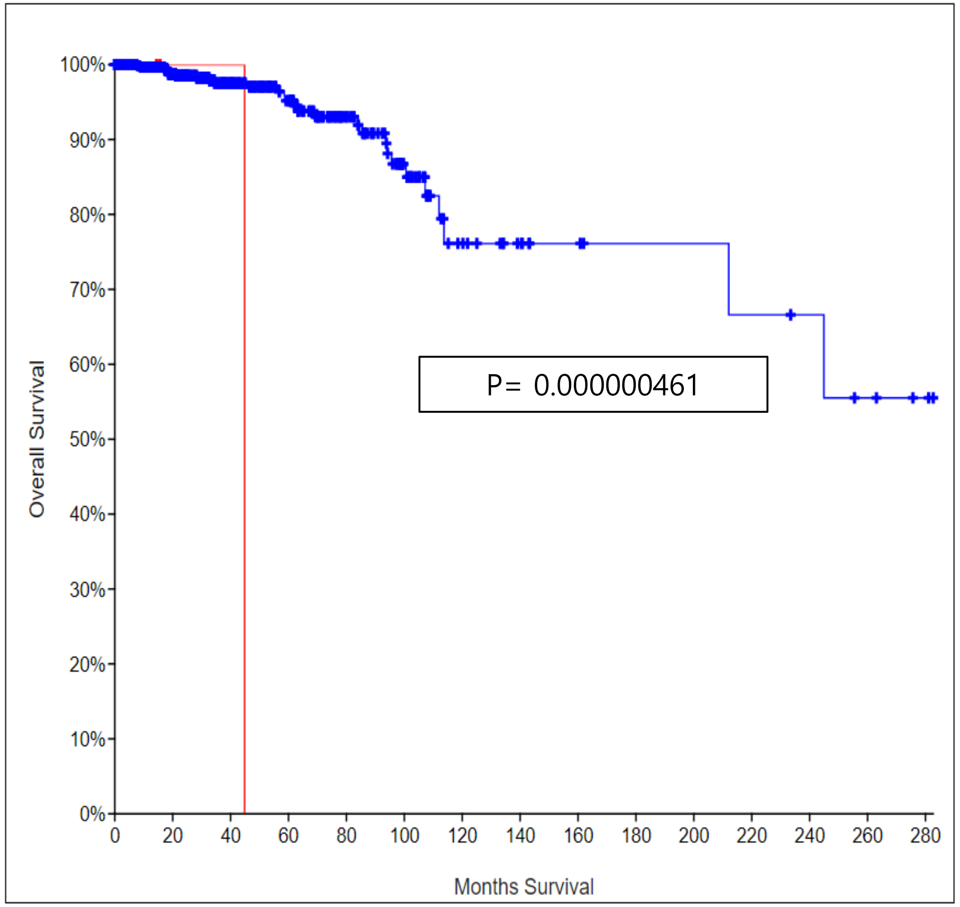


B Disease/Progression-free Kaplan-Meier Estimate

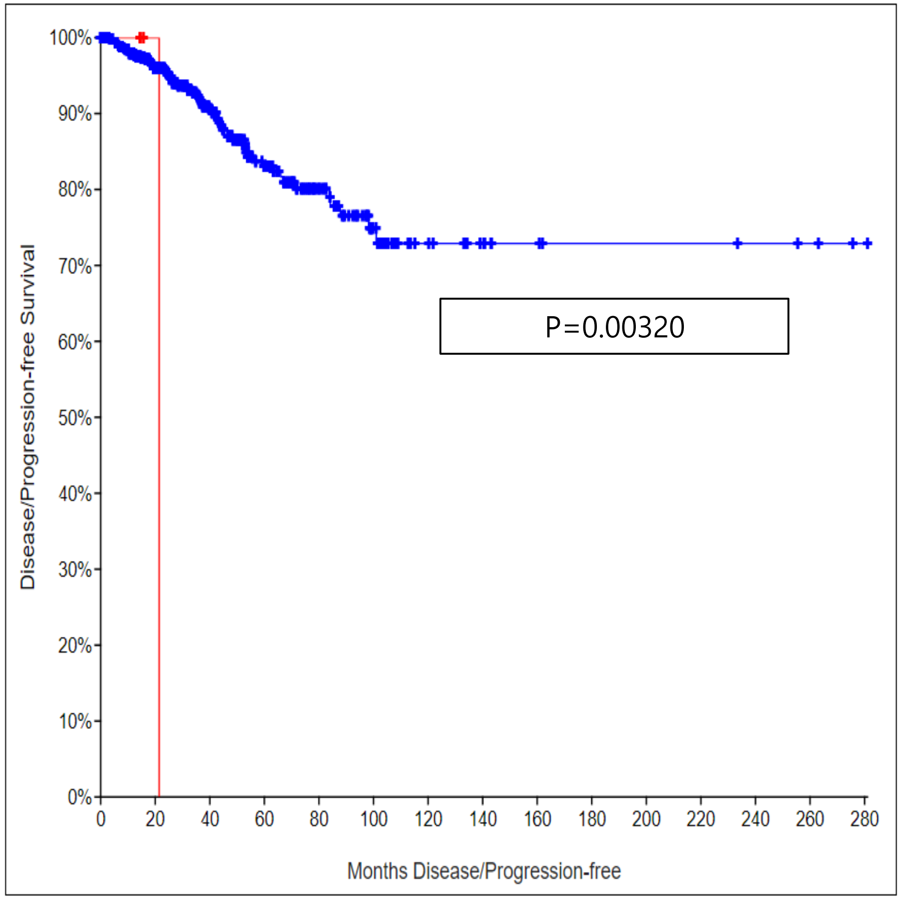


| | Number of Cases, Total | Number of Cases, Deceased | Median Months Survival |
|---|---|---|---|
| Cases with Alteration(s) in Query Gene(s) | 3 | 1 | 44.84 |
| Cases without Alteration(s) in Query Gene(s) | 649 | 28 | NA |

| | Number of Cases, Total | Number of Cases, Relapsed/Progressed | Median Months Disease-free |
|---|---|---|---|
| Cases with Alteration(s) in Query Gene(s) | 3 | 1 | 21.45 |
| Cases without Alteration(s) in Query Gene(s) | 649 | 62 | NA |

### Figure 7. KM-Curve: TPI1 survival specific mutation gene

**TPI1**

A Overall Survival Kaplan-Meier Estimate

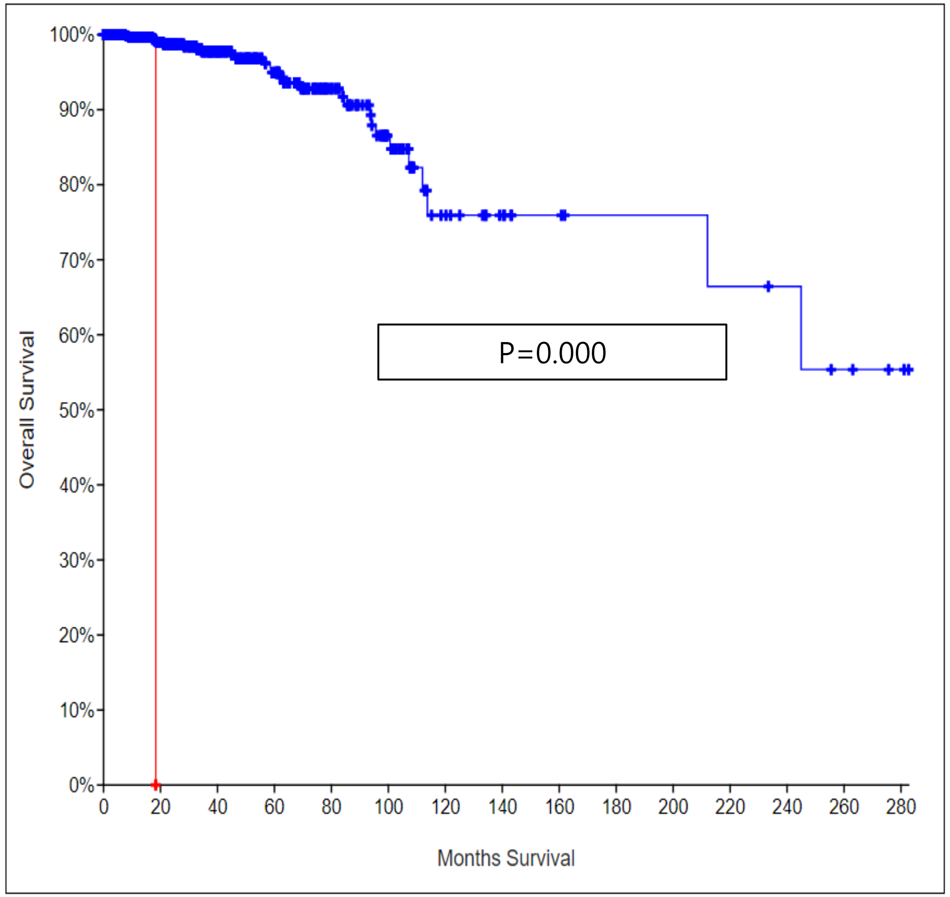


| | Number of Cases, Total | Number of Cases, Deceased | Median Months Survival |
|---|---|---|---|
| Cases with Alteration(s) in Query Gene(s) | 1 | 1 | 18.33 |
| Cases without Alteration(s) in Query Gene(s) | 651 | 28 | NA |

B Disease/Progression-free Kaplan-Meier Estimate

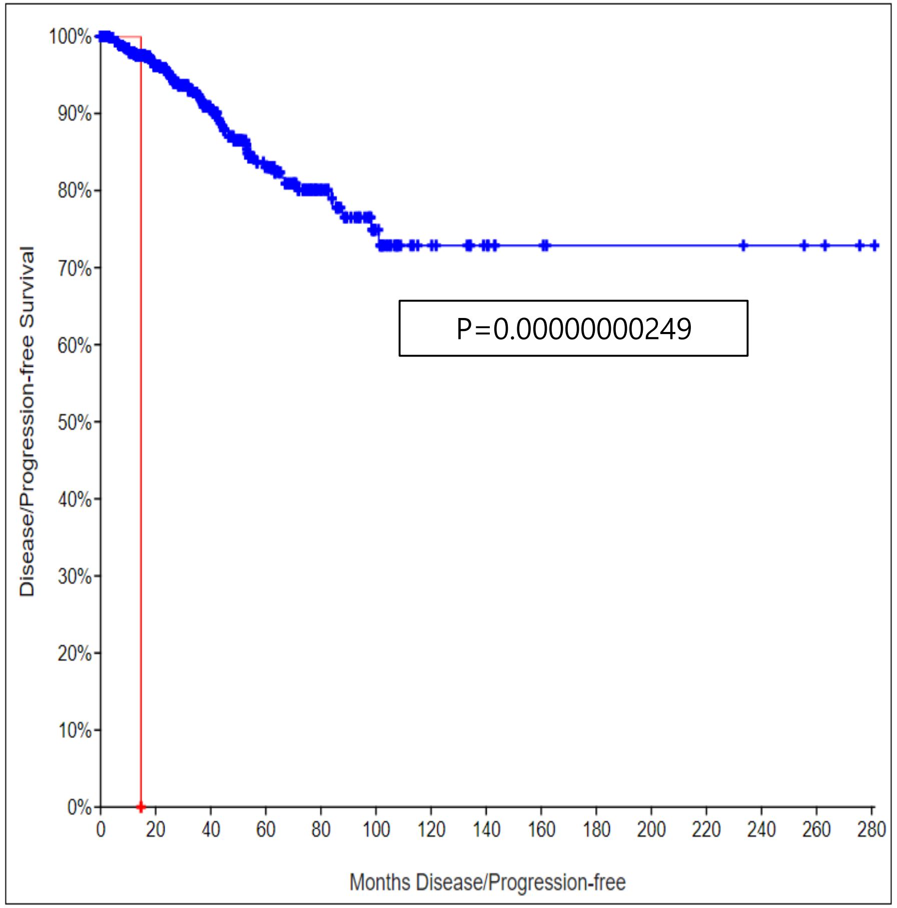


| | Number of Cases, Total | Number of Cases, Relapsed/Progressed | Median Months Disease-free |
|---|---|---|---|
| Cases with Alteration(s) in Query Gene(s) | 1 | 1 | 14.68 |
| Cases without Alteration(s) in Query Gene(s) | 651 | 62 | NA |

**Figure 8. KM-Curve: VWA5B2 survival specific mutation gene**

**VWA5B2**

A Overall Survival Kaplan-Meier Estimate

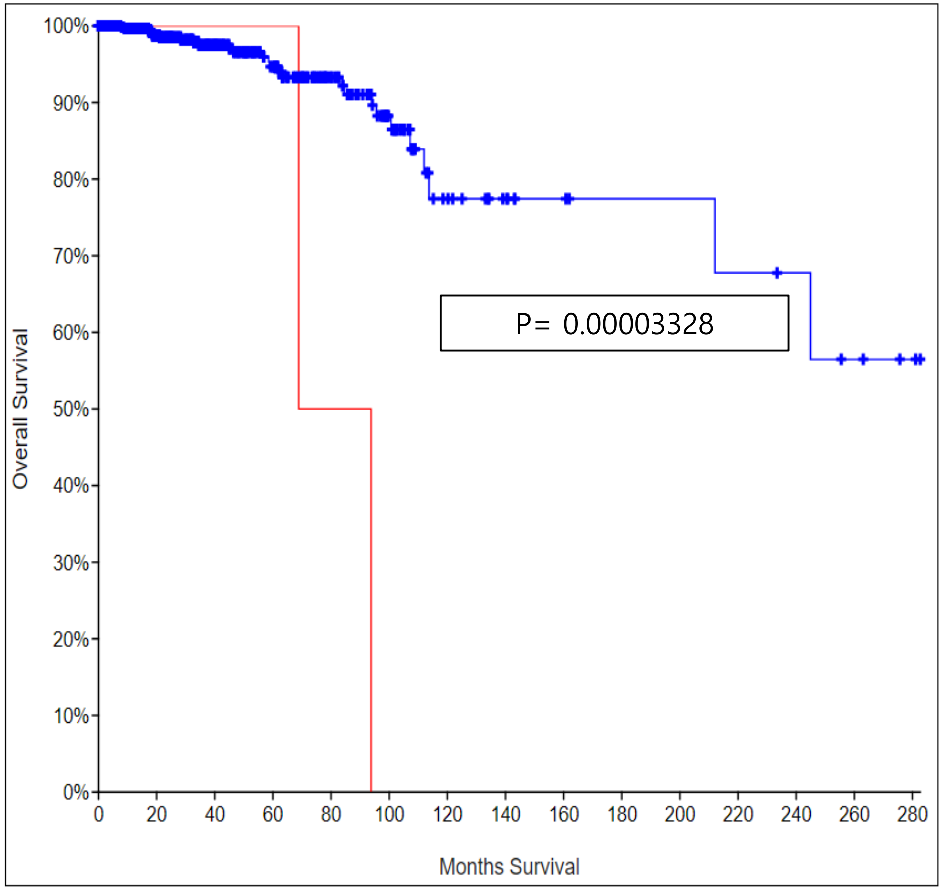


| | Number of Cases, Total | Number of Cases, Deceased | Median Months Survival |
|---|---|---|---|
| Cases with Alteration(s) in Query Gene(s) | 3 | 2 | 68.89 |
| Cases without Alteration(s) in Query Gene(s) | 649 | 27 | NA |

B Disease/Progression-free Kaplan-Meier Estimate

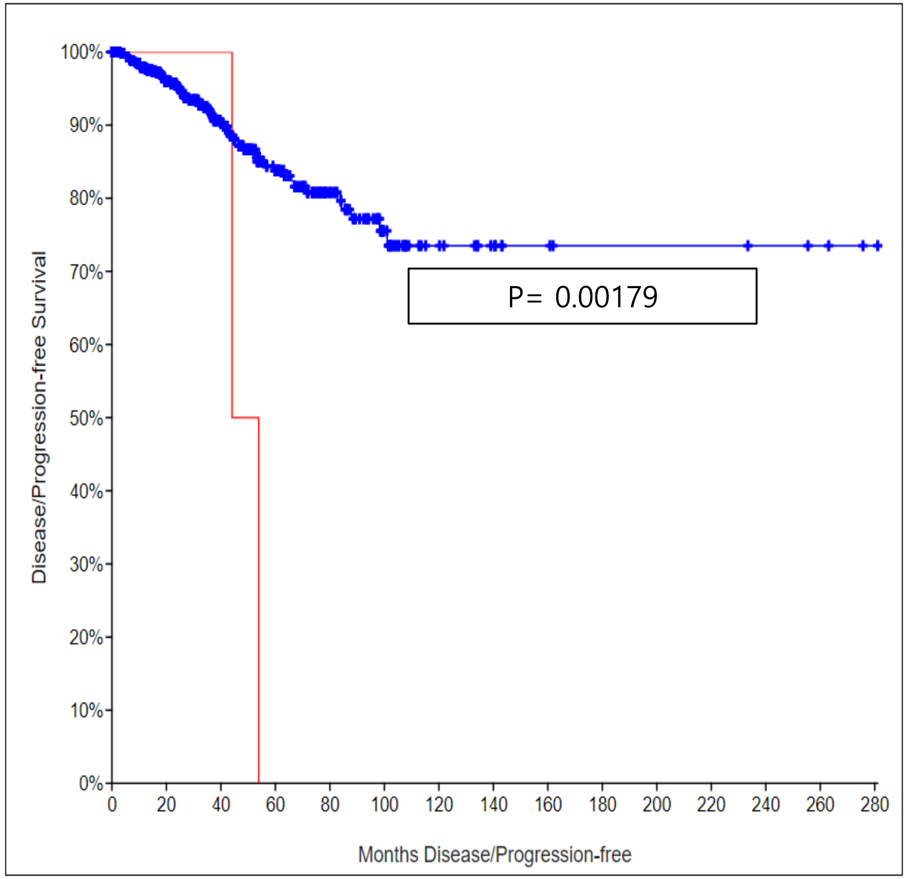


| | Number of Cases, Total | Number of Cases, Relapsed/Progressed | Median Months Disease-free |
|---|---|---|---|
| Cases with Alteration(s) in Query Gene(s) | 3 | 2 | 44.12 |
| Cases without Alteration(s) in Query Gene(s) | 649 | 61 | NA |
